# DPP4 inhibitors and respiratory infection, a systematic review and meta-analysis of the CardioVascular Outcomes Trials conducted before the pandemic and implications for the management of diabetes during COVID-19

**DOI:** 10.1101/2020.07.28.20163386

**Authors:** Guillaume Grenet, Samia Mekhaldi, Sabine Mainbourg, Marine Auffret, Catherine Cornu, Jean-Luc Cracowski, François Gueyffier, Jean-Christophe Lega, Michel Cucherat

## Abstract

**Background:** Association between DPP4 inhibitors and respiratory infection remains unclear. CardioVascular Outcomes Trials (CVOTs) conducted before the COVID-19 pandemic are available. We aimed to estimate the effect of DPP4 inhibitors on the risk of respiratory infections.

**Methods:** We updated a previous systematic review and meta-analysis, searching for CVOTs assessing a DPP4 inhibitor in patients with type 2 diabetes mellitus. We focused on placebo-controlled CVOTs. Our primary outcome was ‘any respiratory infection’. We added a sensitivity analysis integrating non-CVOTs and active-controlled CVOTs.

**Findings:** We included 47 714 patients in five placebo-controlled CVOTs. Median follow-up ranged from 1·5 years to 3 years. 4 369 events of overall respiratory infection were reported (rate of 9·2%). DPP4 inhibitors were not associated with a different risk compared to placebo (RR = 0·99 [95% CI: 0·93; 1·04]). The sensitivity analysis integrating the non-CVOTs studies and the active-controlled CVOT reached 11 349 events among 82 644 participants (rate of 13·7%). DPP4 inhibitors were not associated with a different risk of overall respiratory infection (RR = 1·00 [95% CI: 0·97; 1·03]).

**Interpretation:** Our up-dated meta-analysis provides the most powerful and least biased estimation of the association of DPP4 inhibitors and the risk of overall (non COVID-19) respiratory infection. We did not find any effect of the DPP4 inhibitors on the risk of respiratory infection. Our results support the recently published practical recommendations for the management of diabetes in patients with COVID-19, suggesting that DPP4 inhibitors should not be discontinued regarding the COVID-19 pandemic.

**Funding:** No source of funding

**Panel: Research in context:** *Evidence before this study:* From before the COVID19 pandemic, respiratory infections are considered potential adverse effects of DPP4 inhibitors. Randomized trials assessing DPP4 inhibitors in patients with type 2 diabetes (T2D), their meta-analyses and pharmacovigilance studies reported conflicting results. Since the last meta-analyses assessing the risk of infections with DPP4 inhibitors, powerful cardiovascular outcomes randomized trials (CVOTs) became available. Recent practical recommendations for the management of diabetes during COVID-19 suggested that DPP4 inhibitors could be continued. We updated our previous meta-analysis of CVOTs and focused to the overall risk of respiratory infection associated with DPP4 inhibitors. We searched for published and unpublished CVOTs in Medline, the Cochrane Central Register of Controlled Trials (CENTRAL), and ClinicalTrials.gov, up to January 27, 2020, using key word as “diabetes mellitus”, “hypoglycemic agents”, “glucose control”, “randomized controlled trial”, “cardiovascular diseases”.

*Added value of this study:* We included CVOTs comparing a DPP4 inhibitor versus placebo, in people with T2D, and analysed the risk of respiratory infection with DPP4 inhibitors. We focused on placebo-controlled CVOTs to avoid the pitfalls of small study effect and heterogeneous comparators. We added a sensitivity analysis integrating non-CVOTs and non-placebo CVOTs to challenge our results and to increase the statistical power. Our meta-analysis provides the most powerful and least biased estimation of the association of DPP4 inhibitors and the risk of overall (non COVID-19) respiratory infection. Our analyses integrated 11 349 events of any respiratory infections through 82 644 patients from randomized trials. Our results did not find any association between DPP4 inhibitors use and risk of non-COVID respiratory infections.

*Implications of all the available evidence:* The current COVID-19 pandemic has raised some questions about pros and cons of certain cardiovascular drugs. Our results support the recent practical recommendations for the management of diabetes in patients with COVID-19, suggesting that DPP4 inhibitors should not be discontinued regarding the COVID-19 pandemic.

## Introduction

From before the COVID19 pandemic, respiratory infections are considered to be potential adverse effect of dipeptidyl peptidase-4 (DPP-4) inhibitors.^1,2^ Some randomized clinical trials (RCT) suggested an increased risk of respiratory (and urinary) infections with sitagliptin;^3^ an increased risk of infection with alogliptin ^4^ –as reported in the figure 2 of the meta-analysis by Yang et al –;^5^ and an increased risk of respiratory infection with linagliptin ^6^ –as reported in the figure 5.a of the meta-analysis by Yang et al –.^5^ Previous meta-analyses showed conflicting results. Richter et al suggested an increased risk of all-cause infection with sitagliptin.^7^ Monami et al reported an increased risk of nasopharyngitis with sitagliptin but a decreased risk of other infections with vildagliptin –see their table 5–^8^. Others did not report a class effect.^5,9,10^ The meta-analysis of Yang et al has specifically assessed the risk of any infection with DPP4 inhibitors.^5^ They did not find an increased risk of infection, despite including 74 studies. However, they included only one cardiovascular outcomes trials (CVOT) —SAVOR TIMI 53—,^11^ which weighted 30% of the meta-analysis of overall infection. SAVOR TIMI 53 did not appear in the meta-analysis for respiratory infection.^5^ Since then, at least three other CVOTs assessing a gliptin versus placebo with a large sample size became available.^12–14^ The last meta-analyses of CVOTs did not report such outcomes (respiratory infections) for DPP4 inhibitors.^15,16^ We did not find on-going meta-analysis specific to this question in PROSPERO (April 29, 2020). Pharmacovigilance studies also showed some discrepancies. A disproportionality analysis in the World Health Organization VigiBase reported an increased reporting of upper respiratory tract infections for users of DPP4 inhibitors,^17^ not confirmed in other pharmacoepidemiological studies.^18–21^ Up to now, the potential effect of DPP4 inhibitors on overall respiratory infections remain unclear. In France, adverse events in patients with COVID-19 are already reported as suspected DPP4 inhibitor’s adverse effect, whenever the causality remains unclear.^22^

Recently published practical recommendations for the management of diabetes in patients with COVID-19 suggested that DPP4 inhibitors should not be discontinued.^23^ On another hand, the DPP-4 enzyme could be a potential target for treating coronavirus infections.^24^ We hypothesized that the meta-analysis of the powerful CVOTs would lead to a significantly increase in the statistical power of the analysis. Indeed, the sample sizes of the SAVOR TIMI 53 ^11^ and the TECOS ^13^ CVOTs –16 492 and 14 671, respectively–already surpass the sample size of the meta-analysis of Yang et al for respiratory infection –16 958 under DPP4 inhibitors and 11 939 under control treatment, see table 3 of Yang et al–.^5^ Moreover, using placebo-controlled CVOTs should provide an estimate at lower risk of bias compared to pharmacovigilance studies. We aimed to provide a powerful and low bias update estimate of the effect of DPP4 inhibitors on the overall risk of respiratory infections, allowing testing the recent recommendations for the management of diabetes during COVID-19 pandemic.

## Methods

We focused on respiratory infections in placebo-controlled CVOT assessing a DPP4 inhibitor. We updated a previous systematic review and meta-analysis.^25,26^ The search strategy was previously published.^26^ We updated the searche for CVOTs assessing a DPP4 inhibitor versus a placebo, in type 2 diabetes mellitus, in PubMed and CENTRAL, up to January 27, 2020. We excluded active control as other gliptin, and other hypoglycemic drug, to avoid heterogeneity of control groups and inconsistency issues of indirect comparisons. We excluded specific populations as children and pregnant women.^25^ Two reviewers independently review the bibliographic references (SMe and GG).

For the sensitivity analysis (see under), we i) retained active-controlled CVOT from the bibliographic search and ii) integrated non-CVOTs from the previous meta-analysis of Yang et al.^5^

In line with the previous meta-analysis specifically designed for testing the risk of infection with DPP4 inhibitors of Yang et al,^5^ we looked at the following conditions as ‘any respiratory infections’: influenza, nasopharyngitis, sinusitis, pharyngotonsillitis, pharyngitis, bronchitis, respiratory tract infection, upper respiratory tract infection, lower respiratory tract infection, pneumonia. For each included trial, we extracted data according to the definition used in each trial. In line with Yang et al,^5^ we added up the different kind of respiratory infection. Data of interest were searched for across the articles and on the Clinical Trials Website.^27^ Two reviewers independently extracted the data (GG and SMa). Discrepancies were resolved by consensus. In line with our previous meta-analysis,^26^ we used the Cochrane Collaboration’s tool for assessing risk of bias in RCTs.^28^

Our primary outcome was the treatment effect on the risk of ‘any respiratory infection’. Secondary outcomes were i) any upper respiratory tract infection (influenza, nasopharyngitis, sinusitis, pharyngotonsillitis, pharyngitis, upper respiratory tract infection); ii) any lower respiratory tract infection (bronchitis, lower respiratory tract infection, pneumonia); and each components themselves (influenza, nasopharyngitis, sinusitis, pharyngotonsillitis, pharyngitis, bronchitis, respiratory tract infection, upper respiratory tract infection, lower respiratory tract infection, pneumonia).

Our primary analysis was the meta-analysis of the placebo-controlled CVOTs only, to avoid the risk of “small study effects” ^29^ and the potential heterogeneity of different comparators. We added a sensitivity analysis integrating i) the combined non-CVOTs from the previous meta-analysis of Yang et al,^5^ and ii) active-controlled CVOT (assessing a gliptin versus other hypoglycemic drug, if applicable). In this way, we were able to combine the information of smaller and shorter RCTs already summarized by Yang et al, and the last powerful CVOTs. Such analysis using smaller studies combined and larger trials apart has been successfully used before.^30^

Treatment effect was estimated using risk ratio (RR) and its 95% confidence intervals (95% CI). P-value <0·05 was considered as significant. Analyses were performed using R 3.6.2 (Package [meta], version 4.11-0).^31^ The heterogeneity between studies was assessed using I^2^ test. We used fixed-effect model when I^2^ was <50%, random-effects model otherwise. Reporting of the study was conducted according to the Preferred Reporting Items for Systematic Reviews and Meta-Analyses (PRISMA) statement.^32^ Publication bias was searched for using funnel plot.

The protocol of the present review has not been registered, but the hypothesis, the bibliographic search, the outcome definition and the statistical plan were defined *a priori*, notably using our previous meta-analysis (which was registered in PROSPERO),^26^ and the previous meta-analysis of Yang et al, specifically designed for assessing the risk of infections with DPP4 inhibitors.^5^

## Results

The results refers to the primary objective (placebo-controlled CVOTs), except for the sensitivity analysis of the primary outcome’s section. Preferred Reporting Items for Systematic Reviews and Meta-Analyses (PRISMA) checklist is available in appendix (Appendix p2-3). The update bibliographic search retrieved 762 references. The selection process of the update is presented in figure 1. Five trials with 47 714 patients were included.^11–14,33^ Complementary to the previous systematic review, this update identified the final publication of one trial,^14^ and one early-terminated trial “MK-3102-018”.^33^ The CAROLINA CVOT was not included in the primary analysis but integrated in the sensitivity analysis, as it compared a DPP4 inhibitor versus a sulfonylurea.^34^

**Figure 1.**
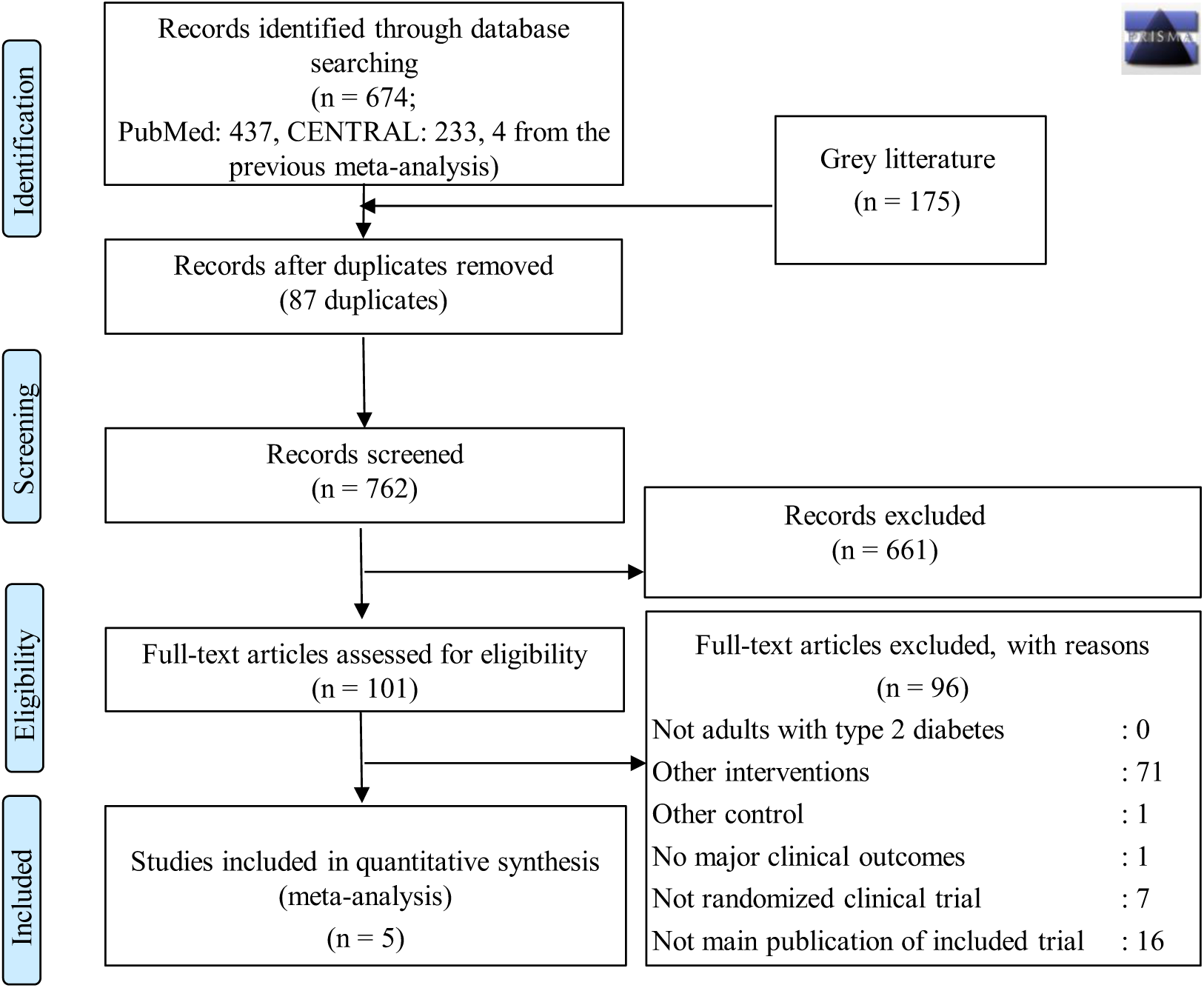
Flow chart of the update of the systematic review restricted to cardiovascular outcomes trials assessing a DPP4 inhibitor versus placebo.

Baseline characteristics of included trials are presented in Table 1. Included trials were published between 2013 and 2019. Median follow-up ranged from 1·5 years to 3 years. Percentage of males ranged from 62·9 to 70·7%, percentage of patients i) with high blood pressure or receiving antihypertensive drugs from 78·4 to 95·4%, ii) with dyslipidemia or receiving statins treatment from 71·2 to 90·4%, iii) receiving antiplatelet treatment from 68·3 to 97·2%, and percentage of current smokers at inclusion from 10·2 to 14·4%. Mean age ranged from 61 to 65·8+/-9·1 years, mean duration of diabetes from around 7·2 to 14·7+/-9·5 years, mean HbA1c at inclusion from 7·2+/-0·5 to 8·01+/-0·87%, mean body mass index (BMI) at inclusion from 28·7 to 31+/-5·3 kg.m^-2^. The risk of bias assessments is presented in appendix (Appendix p4). All studies were randomized double-blinded placebo controlled trials. The risk of reporting bias of respiratory infections was unclear for all the included study. Only sparse data regarding respiratory infection were available for the TECOS trial.^13^ Contacted, the sponsor explained that the safety data were unfortunately not available for safety meta-analysis. Definitions of the events for each trial are presented in appendix (Appendix p5).

**Table 1.**
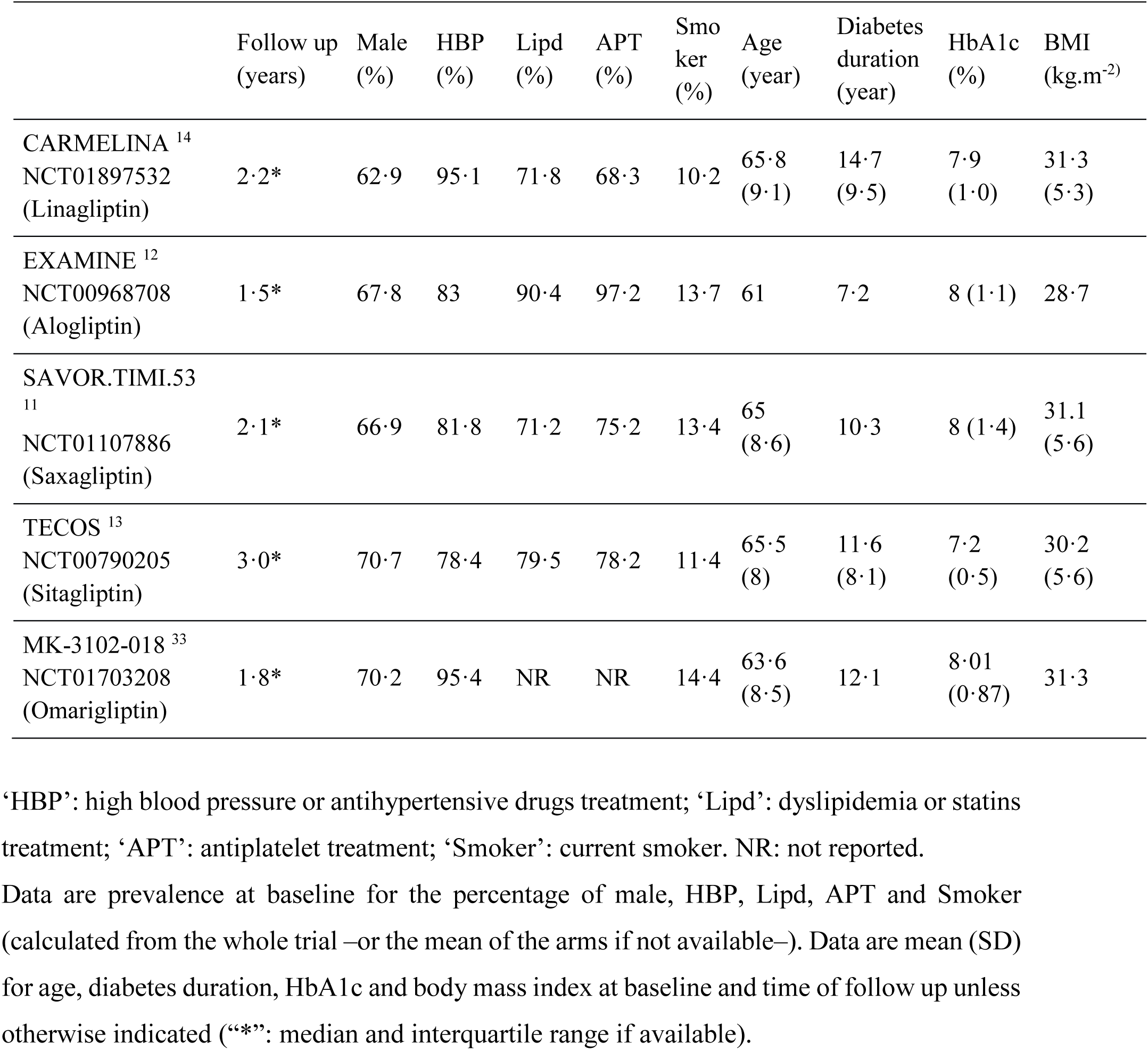
Baseline characteristics of patients in the included trials

### Primary outcome

Five studies contributed to this analysis, including 4 369 events among 47 714 patients (rate of 9·2%). DPP4 inhibitors were not associated with a different risk of overall respiratory infection compared to placebo (RR = 0·99 [95% CI: 0·93; 1·04]). The heterogeneity was low (0%). Forest plot is shown in figure 2. Visual analysis of the funnel plot did not suggest a publication bias (see Appendix page 6).

**Figure 2.**
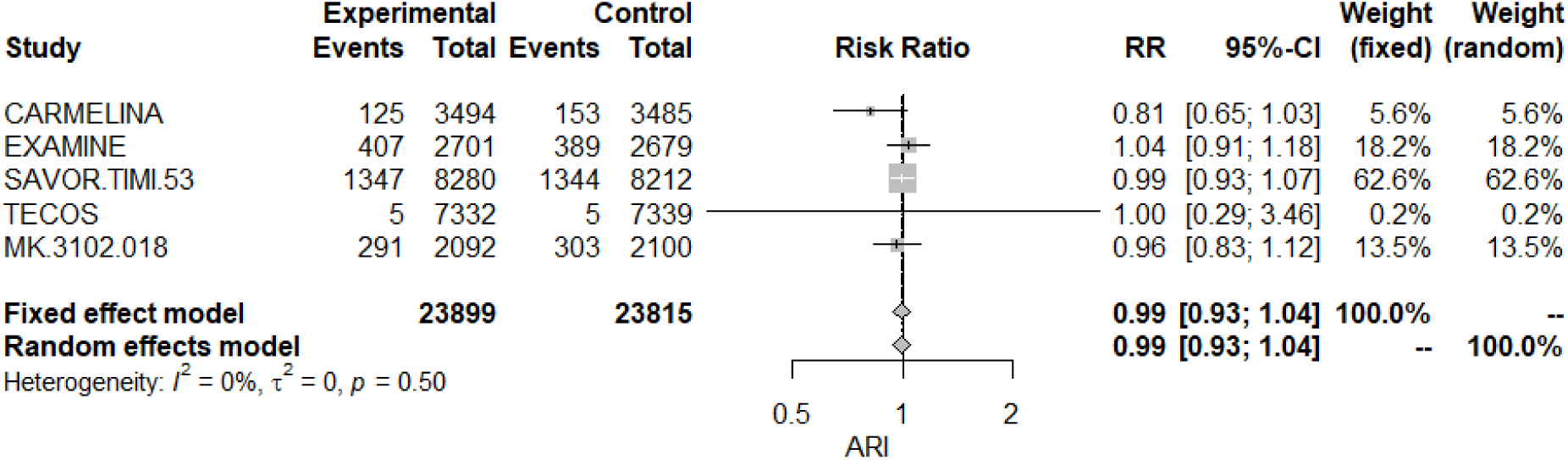
Forest plot for the primary outcome, primary analysis. ARI: any respiratory infection

### Sensitivity analysis of the primary outcome

With the integration of the non-CVOTs studies and of active-controlled CVOT in the meta-analysis, we reached 11 349 events among 82 644 participants (rate of 13·7%). DPP4 inhibitors were not associated with a different risk of overall respiratory infection (RR = 1·00 [95% CI: 0·97; 1·03]). The heterogeneity was null (0%). Forest plot is shown in figure 3.

**Figure 3.**
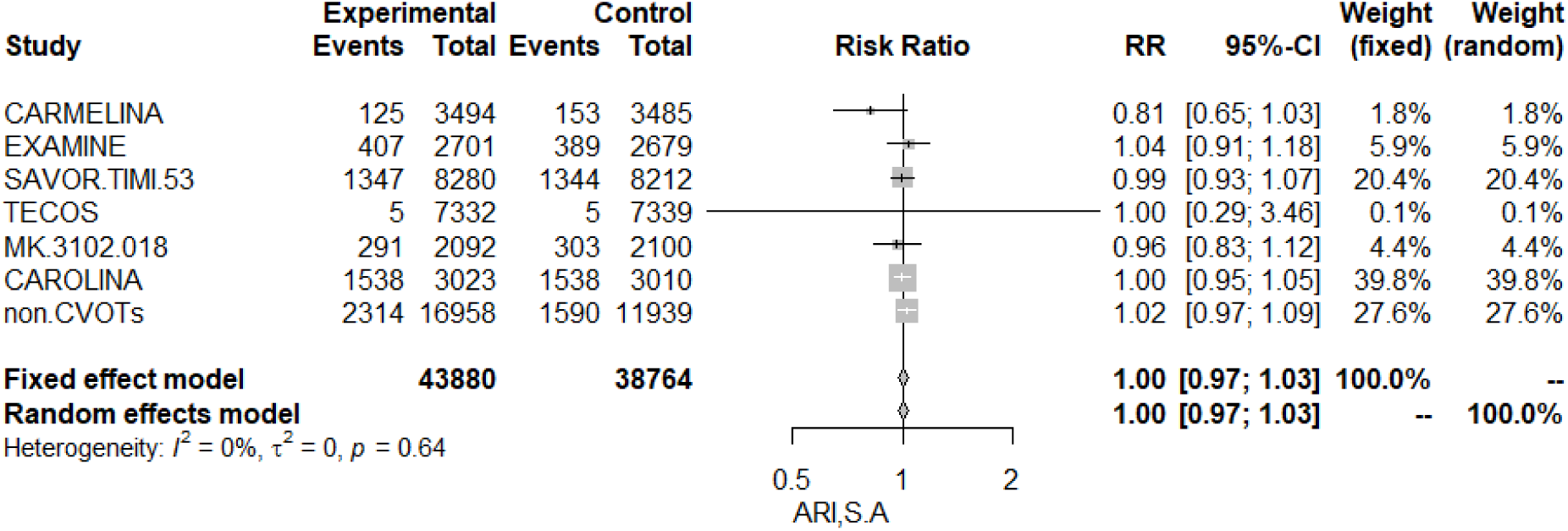
Forest plot for the sensitivity analysis of the primary outcome, integrating the placebo-controlled CVOTs, the active-controlled CVOT (here, the CAROLINA trial), and the non-CVOTs combined from the meta-analysis of Yang et al.^5^ ARI: any respiratory infection; S.A: sensitivity analysis

### Secondary Outcomes (primary analysis)

Five studies contributed to the meta-analysis of any upper respiratory tract infection, including 2 699 events. DPP4 inhibitors were not associated with a different risk of any upper respiratory infection compared to placebo (RR = 1·00 [95% CI: 0·93; 1·08]). Heterogeneity was moderate (I^2^=31%). The forest plot is shown in appendix (Appendix p7). Five studies contributed to the meta-analysis of any lower respiratory tract infection, including 1 650 events. DPP4 inhibitors were not associated with a different risk of any lower respiratory infection compared to placebo (RR = 0·96 [95% CI: 0·87; 1·06]). Heterogeneity was moderate (41%). The forest plot is shown in appendix (Appendix p7). DPP4 inhibitors were not associated with different risks regarding each components of the respiratory infections (forests plots are displayed in appendix, p7-10).

## Discussion

Our up-dated meta-analysis provides the most powerful and least biased estimation of the association of DPP4 inhibitors and the risk of overall (non COVID-19) respiratory infection. We did not find any effect of the DPP4 inhibitors on the risk of respiratory infection. Our results support the recently published practical recommendations for the management of diabetes in patients with COVID-19, suggesting that DPP4 inhibitors should not be discontinued regarding the COVID-19 pandemic.^23^

Our study has strengths and limitations. To the best of our knowledge, our study is the most comprehensive meta-analysis of large CVOTs assessing the risk of respiratory infection with DPP4 inhibitors, in patients with type 2 diabetes, before the COVID-19 pandemic. We even found one early-terminated CVOT not included in the last meta-analysis of glucose lowering drugs.^16^ We summarized the information from five placebo-controlled CVOTs, including 47 714 patients. We firstly focused on placebo-controlled CVOTs, to avoid both the pitfalls of small study effect and of heterogeneous comparators. This could lead to a loss of information. However, the non-CVOTs have a relatively small sample size and time of follow up compared to the CVOTs. Thus, we expected the CVOTs to drive the main number of events. A previous meta-analysis of non-CVOTs DPP4 inhibitors studies provided treatment effect estimates on cardiovascular outcomes eventually disproved by CVOT.^35^ To limit the risk of a loss of information with our methodological choice, we added a sensitivity analysis to verify the treatment effect estimation using both active-controlled CVOTs and non-CVOTs studies, which showed the robustness of our primary analysis. The placebo-controlled CVOTs reported more events (4 369) than the previous meta-analysis of non-CVOTs studies (3 904).^5^ Integrating the placebo-controlled CVOTs, the active-controlled CVOT and the non-CVOTs studies, we provide a very precise estimation using an overall sample size of 11 349 events and 82 644 patients. Finally, we provide an estimation with a lower risk of bias and a greater statistical power compared to the previous pharmacovigilance studies. Indeed, they reported a number of events in patients exposed to DPP4 inhibitors ranging from 25 hospitalizations for community-acquired pneumonia,^18^ 87 pneumonia,^20^ 242 reports of any infections,^17^ 537 acute respiratory infections (compared to sulfonylureas),^19^ and to 2440 infections overall.^21^

We considered the risk of reporting bias as unclear for all the included study in our meta-analysis. Indeed, our outcomes of interest were not prespecified outcomes of interest of the included trials. Moreover, the reporting was heterogeneous between trials. This is especially the case for the TECOS trial, which surprisingly reported a very low rate of respiratory infections, despite a large sample size. However, the risk of reporting bias within each trial (i.e. between groups in each trial) was not considered as high, as most of the adverse events were “collected by systematic assessment” (see their report on the Clinical Trial website), in a double-blinded fashion, and coded using the Medical Dictionary for Regulatory Activities (MedDRA) definitions. Finally, as only one placebo-controlled CVOT was available per DPP4 inhibitor, we cannot exclude a molecule effect.

It should be noted that the present review focused on the risk of respiratory infections and DPP4 inhibitors. The absence of demonstrated cardiovascular benefits of DPP4 inhibitors use in patients with type 2 diabetes, contrary to glucagon-like peptide-1 (GLP1) agonist receptors and Sodium-glucose Cotransporter-2 (SGLT2) inhibitors, has been documented before.^15,26^ The question of the role of the DPP4 inhibitors in type 2 diabetes management in general is beyond the scope of this paper.

## Conclusion

The current COVID-19 pandemic has raised many questions, especially regarding type 2 diabetes management.^23^ We did not find any association between DPP4 inhibitors use and the risk of non-COVID respiratory infection. On-going randomized trial assessing sitagliptin in COVID-19 positive patients will help further understand the relationship between DPP4-inhibitors and COVID-19 infections.^36^

## Data Availability

Data are available in the forest plots of the appendix.

## Abbreviations

AE: Adverse Events
95% CI: 95% Confidence Intervals
ALRTI: Any Lower Respiratory Tract Infection
APT: Antiplatelet Treatment
ARI: Any Respiratory Infection
AURTI: Any Upper Respiratory Tract Infection
BMI: Body Mass Index
CENTRAL: Cochrane Central Register of Controlled Trials
CVOT: CardioVascular Outcomes Trials
DPP-4: DiPeptidyl Peptidase-4
GLP1: Glucagon-Like Peptide-1
HbA1c: Glycated Hemoglobin
HBP: High Blood Pressure or antihypertensive drugs treatment
Lipd: Dyslipidemia or statins treatment
LRTI: Lower Respiratory Tract Infection
MedDRA: Medical Dictionary for Regulatory Activities
NR: Not Reported
PRISMA: Preferred Reporting Items for Systematic Reviews and Meta-Analyses
RCT: Randomized Clinical Trials
RR: Risk Ratio
RTI: Respiratory Tract Infection
S.A: Sensitivity Analysis
SAE: Serious Adverse Events
SD: Standard Deviation
SGLT2: Sodium-Glucose Cotransporter-2
Smoker: Current smoker
T2D: Type 2 Diabetes
URTI: Upper Respiratory Tract Infection

## Contributors

GG and SMe did the literature search, GG and SMa the data extraction. GG conducted the analyses under the supervision of MCu. GG wrote the first draft of the report. SMa, MA, CC, JLC, FG, JCL and MCu contributed to data interpretation of the findings and provided critical input for important intellectual content.

## Declaration of interests

GG, SMe, SMa, MA, CC, JLC, declare that they have no competing interest.

JLC reports investigator driven grants from Bioprojet Pharma, Pfizer and United Therapeutics outside the submitted work.

JCL has received speaking fees and honoraria from Roche.

MC has received consulting fees from Boehringer Ingelheim, SANOFI, AstraZeneca and speaker honoraria from SANOFI.

In the last five years, FG received for his institution fees from Portola Pharmaceuticals for central reading of ultrasound records, from Neurochlore for DSMB coordination, from EryTech Pharma for modeling projects, from RCTs and Steve Consultant for exploring French social security database.

Ethics approval and consent to participate: not applicable.

